# EpiCurveBench: Evaluating epidemic curve digitization

**DOI:** 10.1101/2025.09.23.25336494

**Authors:** Thomas Berkane, Maimuna S. Majumder

## Abstract

Accurate data on disease case counts over time is essential for training reliable disease forecasting models. However, such data is often locked in non-machine-readable formats, most commonly as epidemic curve (epicurve) images—charts that depict case counts of a given disease over time, for a given location. Digitizing these charts would greatly expand the data available for forecasting models, improving their accuracy. Manual digitization, though, is very time-consuming, and existing automated methods struggle with real-world epicurves due to dense datapoints, overlapping series, and varied visual styles. To address this, we present EpiCurveBench, a benchmark of 100 manually curated and annotated epicurve images collected from diverse sources. The dataset spans a wide range of chart styles, from simple to highly complex. We also introduce Epi-Curve Similarity (ECS), a new evaluation metric that captures the temporal structure of epicurves, handles series of varying lengths, and remains stable in the presence of incomplete data. Using this metric, we evaluate state-of-the-art chart data extraction methods on EpiCurveBench and find substantial room for improvement, with the best model achieving an ECS of only 42.9%. We release the dataset and evaluation pipeline to accelerate progress in epicurve extraction. More broadly, the difficulty of EpiCurveBench compared to existing chart extraction benchmarks provides a rigorous testbed for advancing chart data extraction methods beyond disease forecasting.

**Institutional Review Board (IRB):** This research does not require IRB approval.

## 1. Introduction

Models that forecast disease case counts over time can reduce disease burden (Brooks et al., 2015), but require historical temporal case data, often sourced from disease outbreak reports. Much of this data exists only as epidemic curve (epicurve) images—charts showing case counts over time for a given disease in a given location—and is thus effectively “locked away.” Epicurves are commonly produced by health agencies in regions experiencing endemicity or epidemics, but their non–machine-readable format limits their use for training forecasting models that guide outbreak response, surveillance, and resource allocation. As a result, public health officials are often asked to compile and format such data on demand—a burdensome, inefficient process—highlighting the need for scalable methods to automatically extract and structure epicurve data.

Improved access to historical epicurve data could directly strengthen epidemic forecasting models, which rely on high-quality time-series data for training and calibration. Unlocking data currently trapped in image form could help enable earlier detection of outbreak trends, better situational awareness, and more informed public-health decision making.

Currently, manual tools like WebPlotDigitizer (Rohatgi) are used to digitize epicurves, but extracting data from a single complex image can take hours. Automated chart data extraction aims to recover the underlying numerical data from chart images. Early methods relied on traditional computer vision and optical character recognition (Farahani et al., 2023). Recently, large vision-language models have achieved stronger performance (Han et al., 2023; Xia et al., 2025). However, these models still struggle with complex, real-world charts. Most existing datasets (Methani et al., 2020; Masry et al., 2022; Xia et al., 2025) consist of synthetic charts with low variety. A notable exception, ChartQA (Masry et al., 2022), includes real-world charts, but they come from only four platforms and are generally simple—basic infographics, pie charts, or bar charts with a low volume of datapoints and clearly printed numbers. The lack of varied, challenging benchmarks has limited progress on more robust extraction methods.

To address this, we introduce EpiCurveBench, a benchmark designed to advance automated epicurve data extraction and benefit the broader chart data extraction community. EpiCurveBench contains 100 carefully curated, hand-annotated epicurve images from diverse sources, spanning a wide range of diseases, locations, time periods, and styles. It includes 30 simpler epicurves for a smooth transition into the task and 70 challenging ones selected for variety and difficulty. These test a data extraction method’s ability to generalize, with many featuring unique visual formats. In particular, extracting data from the challenging subset requires fine-grained visual understanding and adaptability, as epicurves often include dense datapoints, overlapping series, rotated or tiny text, low-resolution images, thin bars, low contrast, and overlaid text. Even human annotators found the task difficult: some epicurves required up to three hours to digitize using tools like WebPlotDigitizer.

Existing evaluation metrics for chart data extraction are poorly suited for epicurves, as they ignore temporal structure, overpenalize small shifts, and fail to distinguish missing values from incorrect ones. To address this, we introduce EpiCurve Similarity (ECS), a measure based on Edit Distance with Real Penalty (Chen and Ng, 2004), which aligns two time series through minimal-cost matching while explicitly handling gaps and temporal misalignments, critical for accurate epicurve evaluation.

We evaluate multiple state-of-the-art automated chart data extraction methods, including frontier vision-language models, on EpiCurveBench using this new metric. Results show that EpiCurveBench is highly challenging: current state-of-the-art methods leave substantial room for improvement, unlike other chart benchmarks that are nearing saturation. EpiCurveBench highlights significant gaps in current methods and lays the groundwork for improving both epidemic forecasting and chart data extraction, advancing visual reasoning in foundation models.

### Contributions

(1) EpiCurveBench, a challenging, real-world benchmark for epicurve data extraction. (2) ECS, a metric tailored to time series extraction. (3) A comprehensive evaluation of state-of-the-art methods. (4) Open-source data and evaluation code to support development of new methods.

## 2. EpiCurveBench

### 2.1 Data collection

We collect epicurve images from diverse sources to capture a wide range of styles and formats. We crawl directory pages from organizations such as the World Health Organization (WHO) regional offices, the Pan American Health Organization (PAHO), and the US Centers for Disease Control and Prevention (CDC), scraping images from webpages and linked outbreak report PDFs. We also run targeted Google Image searches to capture additional images.

This yields 1,397 epicurve images, from which we manually curate 100 that are openly licensed, prioritizing those that are most varied and challenging. These form two subsets: 30 Basic epicurves (no more than two series per chart and *≤* 30 points per series) and 70 Advanced epicurves with maximal stylistic variety and difficulty. The Basic subset provides an easier entry point for method development.

Appendix A shows the distribution of sources for the curated dataset.

### 2.2 Data annotation

We manually annotate all epicurve images using WebPlotDigitizer. After calibrating both axes with two known points, the annotator clicks each datapoint to record its x and y values. Each epicurve yields a CSV with x-axis values and one or more columns for series values, using original axis labels and series names for consistency. When multiple series are present, we extract all of them, including any non-case-count data such as deaths or rainfall. Appendix B describes how ambiguous cases are handled, and the same instructions are given to evaluated models for consistency between human and automated outputs.

A single annotator completes the initial annotations. To assess reliability, a second annotator independently annotates 20 randomly selected curves from the 70 Advanced epicurves, yielding an inter-annotator ECS of 94.51% (see Section 2.4).

Annotation times range from 2 to 179 minutes, with a mean of 23.7 minutes, a median of 16 minutes, and a standard deviation of 24.7 minutes.

### 2.3 Dataset characteristics

Figure 1 shows sample epicurves. The first row contains three examples from the Basic subset, while the remaining rows show Advanced epicurves, illustrating varied styles and formats. Challenges include thin bars, low resolution, heavy text overlays, stacked bars with similar colors, and lower-resolution historical plots with sub-legible values.

**Figure 1.**
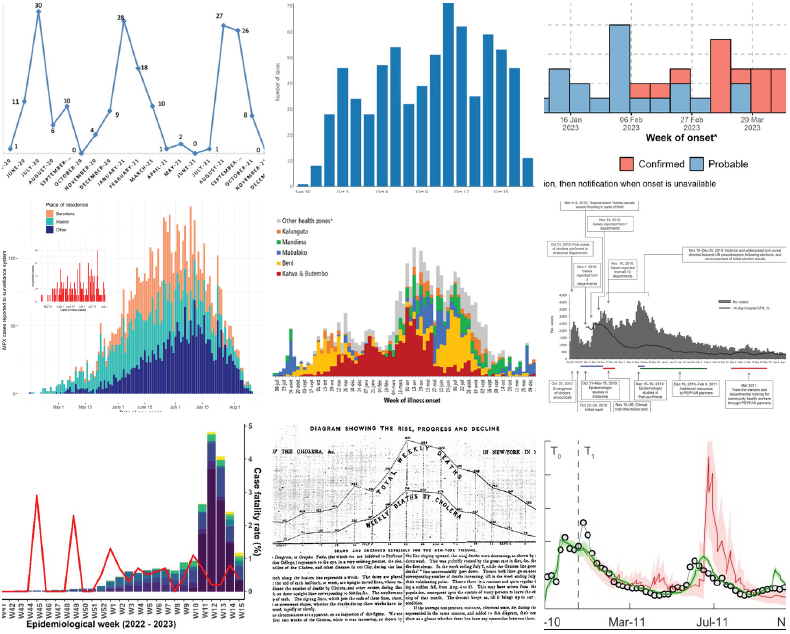
Sample images from EpiCurveBench. Parts of the images, including axes, are truncated for space.

The benchmark includes 57 bar charts, 14 line charts, and 29 mixed charts. Series per image range from 1 to 14 (mean 2.8, median 2, SD 2.4). Series lengths range from 5 to 742 points (mean 95, median 50, SD 130). Charts span 36 countries and the years 1849–2025. Image widths range from 334–1650 pixels and heights from 148–1434 pixels. This variety of chart types, time periods, and visual styles makes EpiCurveBench especially challenging, requiring models to generalize across highly variable conditions for robust performance.

### 2.4 Metric

Two metrics are commonly used for evaluating chart data extraction: Relative Mapping Similarity (Liu et al., 2023, RMS) and Structuring Chart-oriented Representation Metric (Xia et al., 2024, SCRM).

We depart from these metrics because they are poorly suited for evaluating epicurve extraction. Both treat extracted points as unordered mappings between x-axis labels and their corresponding values, thereby ignoring the temporal structure of an epicurve, i.e., case counts over time. Indeed, extracted curves often exhibit temporal shifts between x-axis labels and series values (e.g., values are correct but labels are offset by a few days, causing misalignment), or contain gaps relative to the ground truth (e.g., missing the first or last few points of the curve). Under existing metrics, such misalignments are penalized as harshly as completely incorrect extractions, since the x-axis labels and values no longer align exactly. For example, a curve shifted by only a few days would be judged as being entirely wrong, even though it preserves the overall temporal trend. In practice, forecasters can often realign such curves with minimal preprocessing, meaning that a slight temporal offset still yields a usable signal.

A more appropriate evaluation should accommodate local shifts and missing values, penalizing them while still recognizing partial temporal correspondence. Moreover, existing metrics typically align extracted and ground-truth x-axis labels using textual similarity. This is problematic because epicurve axes are often sparsely labeled, and mismatches can—for instance—occur simply due to differences in date formats. Robust evaluation therefore requires metrics that can handle temporal misalignments and differing series lengths without depending on text-based matching of x-axis labels.

To address these limitations, we introduce Epi-Curve Similarity (ECS), a metric specifically designed for evaluating epicurve extraction. ECS builds on Edit Distance with Real Penalty (Chen and Ng, 2004, ERP), which uses dynamic programming to align series while accounting for gaps and unmatched regions. Like ERP, ECS supports insertions, deletions, and substitutions, and applies real-valued penalties to quantify differences between matched points.

Since epicurve images often contain multiple time series, we first match each automatically extracted series (extraction methods are introduced in Section 3.1) to its ground-truth, annotated counterpart. Let *E* = *{e*_1_, …, *e*_*m*_*}* denote the set of extracted series from an image and *T* = *{t*_1_, …, *t*_*n*_ *}* the corresponding set of ground-truth series for that image. Following Liu et al. (2023); Furkan Biten et al. (2019), we perform matching using the Normalized Levenshtein Distance (NLD), which measures textual similarity (ranging from 0 to 1) between the labels of extracted and ground-truth series (e.g., from chart legends).Two series are matched if their NLD is greater than 0.5. Extracted series with no matching ground-truth are ignored, while unmatched ground-truth series are assigned a score of zero.

For each matched pair of series *p* = (*p*_1_, …, *p*_*P*_) and *t* = (*t*_1_, …, *t*_*T*_), where *i* and *j* index values in the predicted and ground-truth series respectively, the ERP distance is computed by evaluating the following recurrence relation for 0 ≤ *i* ≤ *P* and 0 ≤ *j* ≤ *T*, using dynamic programming:

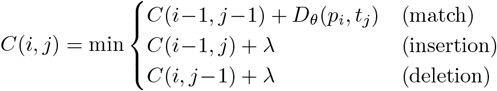

with boundary conditions *C*(0, 0) = 0, *C*(*i*, 0) = *i · λ*, and *C*(0, *j*) = *j · λ*. We fix the gap penalty at *λ* = 1 (as in Liu et al. (2023)).

Intuitively, ERP aligns two series by finding the least-cost path that makes them match. At each step (*i, j*), it decides whether to: (1) match the two points (*p*_*i*_, *t*_*j*_) and pay a cost for their difference; (2) skip a point in the predicted series (an insertion) with a fixed cost *λ*; or (3) skip a point in the ground-truth series (a deletion) with a fixed cost *λ*. The algorithm fills a dynamic programming grid with these cumulative costs and selects the minimum-cost path over all possibilities, thereby measuring the similarity between two curves while allowing for gaps and local temporal shifts.

The distance between two matched points is defined using a relative distance function, normalized by the range of the series’ y-axis:

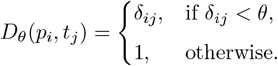

Where

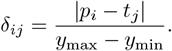

Here, *δ*_*ij*_ denotes the relative difference between the predicted value *p*_*i*_ and the target value *t*_*j*_. *y*_max_ and *y*_min_ denote respectively the maximum and minimum values on the y-axis from which the series values are read, and are collected during the annotation process. The threshold *θ* = 0.01 controls sensitivity: we only tolerate discrepancies within this margin. That is, differences smaller than 1% of the y-axis magnitude are penalized proportionally to their relative magnitude, whereas larger differences are treated as entirely incorrect and assigned the maximum distance of 1.

The ERP distance between two series is then *C*(*P, T*), representing the minimum cumulative cost of transforming *p* into *t*.

The EpiCurve Similarity (ECS) metric normalizes this cost to yield a similarity score between 0 and 1:

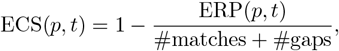

where #gaps = #insertions + #deletions.

For each image, we compute the mean ECS across all matched series, assigning a score of zero to any unmatched ground-truth series. The final performance measure is then obtained by averaging these image-level scores across the entire dataset.

## 3. Baseline performance

### 3.1. Methods evaluated

We evaluate several methods on EpiCurveBench, including two state-of-the-art vision-language models (VLMs) specialized in chart data extraction: OneChart (Chen et al., 2024) and TinyChart (Zhang et al., 2024). We also test three frontier VLMs: GPT-5, Claude Sonnet 4.5, and Gemini 2.5 Pro, each under three configurations: minimal reasoning effort, high reasoning effort, and high reasoning effort with access to a code interpreter tool. For frontier VLMs, we use the prompt in Appendix C.

### 3.2. Results

Table 1 reports ECS scores for both the Basic and Advanced epicurve subsets and the total inference costs for processing the 100 images.

**Table 1:** Performance of all evaluated methods on EpiCurveBench. Results are reported as ECS scores (%) for the Basic and Advanced subsets, along with total inference costs (USD) for the 100 images.

| Model | Basic (%) | Advanced (%) | Cost |
| --- | --- | --- | --- |
| OneChart | 40.3 | 5.8 | — |
| TinyChart | 39.9 | 8.6 | — |
| GPT-5 Minimal | 58.6 | 28.0 | \$0.85 |
| GPT-5 High | 55.6 | 22.4 | \$8.50 |
| GPT-5 High+Code | 58.0 | 24.1 | \$8.13 |
| Claude Minimal | 68.5 | 33.0 | \$1.54 |
| Claude High | 65.4 | 35.5 | \$8.43 |
| Claude High+Code | 64.5 | 34.5 | \$30.12 |
| Gemini Minimal | **70.2** | 40.9 | \$2.56 |
| Gemini High | 68.1 | **42.9** | \$7.24 |
| Gemini High+Code | 67.9 | 35.9 | \$9.17 |

The results show that EpiCurveBench is challenging for all tested approaches. The best-performing models reach an ECS of only 70.2% on the Basic subset (Gemini 2.5 Pro Minimal) and 42.9% on the Advanced subset (Gemini 2.5 Pro High).

OneChart frequently misreads series or axis labels, limiting performance, while TinyChart handles labels better but still produces inaccurate values.

Among frontier VLMs, Gemini 2.5 Pro performs best on both subsets, followed by Claude Sonnet 4.5 and then GPT-5. Across models, the minimal reasoning configuration performs unexpectedly well, outperforming other variants on the Basic subset and yielding the best Advanced subset performance for GPT-5. This suggests that limiting reasoning steps can sometimes prevent errors introduced by reasoning. Adding the code interpreter has mixed effects: it slightly improves GPT-5 while reducing cost, but reduces accuracy and increases cost for other models.

Figure 2 shows the breakdown of error types across frontier VLMs. Numerical errors—imprecise extraction of series values—are the most common. Surplus datapoints (insertions), missed datapoints (deletions), label mismatches, and missed series are less frequent overall but more common in the Advanced set. Notably, missed datapoints often arise when models extract at a coarser temporal resolution than the chart. For the Advanced set, all models, especially under High reasoning effort, show a notable rate of refusals, where the model declines to extract data because it is insufficiently confident. This behavior is most pronounced for GPT-5. The prevalence of numerical errors highlights a broader limitation of multimodal models: difficulty distinguishing fine-grained visual elements (Liu et al., 2025; Razeghi et al., 2024), making dense time-series extraction an open challenge.

**Figure 2.**
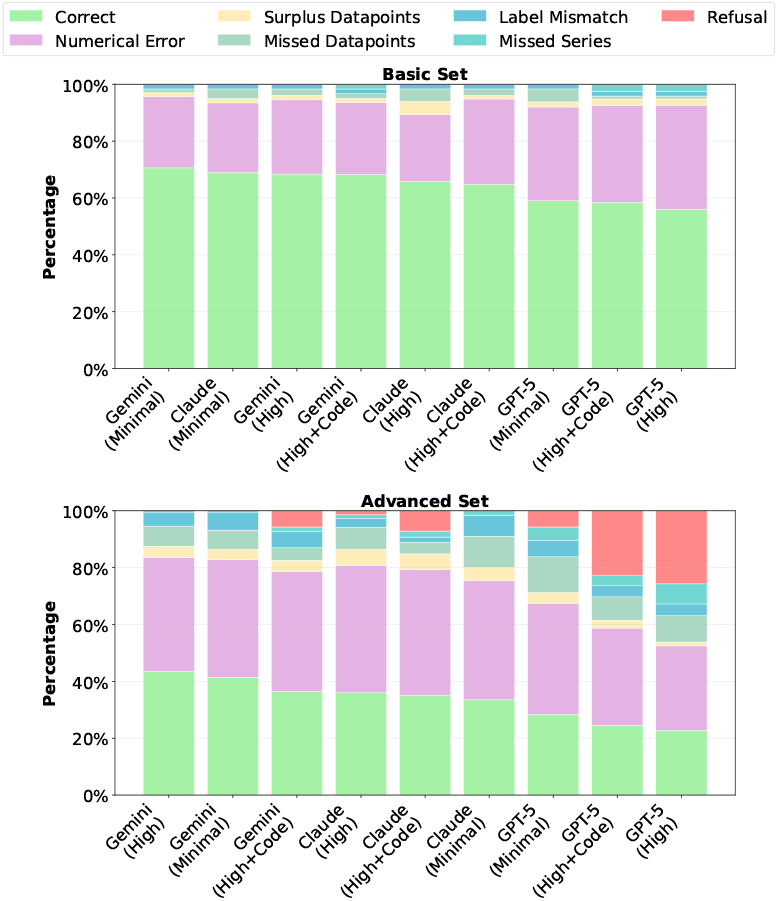
Error types observed during epicurve extraction by frontier VLMs. Percentages indicate the proportion of total ECS loss attributable to each error type. *Numerical Error* : distance between matched points; *Surplus Datapoints* : insertions in the predicted series; *Missed Datapoints*: deletions in the predicted series; *Label Mismatch*: extracted series label does not correspond to any ground-truth (GT) label; *Missed Series*: series present in the GT but not extracted; *Refusal* : model declined to extract.

**Figure 3.**
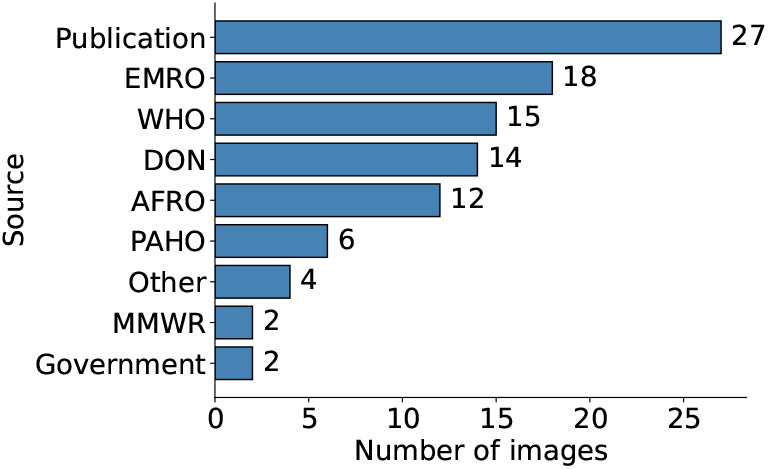
Distribution of EpiCurveBench sources by origin. There are 12 unique source types. The “Other” category includes sources that appear only once. Most publication sources come from ScienceDirect and ResearchGate.

## 4. Limitations and future work

EpiCurveBench currently includes only 100 manually annotated images, limiting the diversity of epicurve formats, and annotations were produced by a single annotator, introducing potential labeling noise. We plan to expand the dataset in future work. Further, we plan to evaluate the downstream impact of extracting data from historical epicurves on epidemic forecasting. For example, CDC Emerging Infectious Diseases reports include epicurves dating back to 1995. Digitizing these historical curves will allow us to quantify how extraction quality affects retrospective forecasting performance, a central task in public health preparedness. Finally, we see potential for extending our framework beyond epicurves to other kinds of health-related charts, such as histograms of chronic illness prevalence and biometric time series.

## 5. Conclusion

We introduce EpiCurveBench, a challenging benchmark for epicurve extraction, along with a new metric for extraction evaluation and an open-source evaluation pipeline. Our evaluation of state-of-the-art models reveals a significant gap between current methods and the complexity of real-world epicurves. We aim to drive the development of generalizable extraction methods to support better epidemic forecasting.

## Data Availability

All data produced in the present study are available upon reasonable request to the authors

## Data and Code Availability

We collect openly licensed epidemic curve images from the web and annotate them. All images, annotations, and metadata are publicly available at https://huggingface.co/datasets/tberkane/EpiCurveBench, and accompanying code is released at https://github.com/tberkane/EpiCurveBench.

## Acknowledgments

We thank our annotator for their careful and time-consuming work in digitizing the epidemic curves. This work was supported by grant IIS2229881 from the National Science Foundation; grant R35GM146974 from the National Institute of General Medical Sciences, National Institutes of Health; and a Moderna Fellowship Award. The funding sources had no involvement in the study design; in the collection, analysis, and interpretation of data; in the writing of the report; or in the decision to submit the paper for publication.

## Appendix A. Distribution of sources

## Appendix B. Handling Annotation Ambiguities

- Only data that is visually present and clearly labeled is annotated. No assumptions are made about missing or implied data.
- Each visible segment in a stacked bar chart is annotated as a separate time series if the figure includes distinct segment labels in the legend or directly on the chart.
- If segment labels are missing, incomplete, or visually indistinguishable, only the aggregate total (height of the full stack) is annotated.

## Appendix C. Frontier VLM Prompt

Here is an image of a chart. Please extract the numerical data it represents and return it in CSV format with appropriate headers. Copy the headers exactly as they are in the image. Make sure to extract data for all x axis values, even those not written on the image, without leaving any gaps. Remember: The sole output should be the CSV table. Nothing else.

